# Cardiac positronium lifetime in human PET: a reproducible right–left ventricular contrast that is not explained by blood oxygenation

**DOI:** 10.64898/2026.06.14.26355630

**Authors:** Ernest Darell Zermeño

**Affiliations:** Universidad Panamericana, Mexico; Universidad de Guadalajara, Guadalajara, Mexico

**Keywords:** positronium lifetime imaging, ortho-positronium, PET/CT, tissue oxygenation, hypoxia biomarker, long axial field of view, confound control

## Abstract

**Background:** Ortho-positronium (o-Ps) lifetime, now measurable in vivo on long-axial-field-of-view (LAFOV) PET/CT, has been proposed as a biomarker of tissue oxygenation and hypoxia. Because o-Ps lifetime is dominated by tissue free-volume structure while the oxygen-specific contribution is small, whether an in-vivo lifetime contrast reflects oxygenation rather than anatomy is an open, identifiability-limited question.

**Aim:** To test the oxygenation hypothesis directly using the heart’s natural arterial/venous oxygenation contrast, with a built-in anatomical control.

**Methods:** We re-analysed a public [^82^Rb]Cl human cardiac LAFOV PET/CT dataset (5.30 × 10^8^ evaluated three-photon events). Per-compartment o-Ps lifetimes were extracted with a background-plus-two-component exponentially-modified-Gaussian (EMG) model. The list-mode→image mapping and right/left ventricle (RV/LV) identity were established lifetime-free (the mapping reproduces the provider’s reconstructed image at block-correlation 0.998 and wins a joint multi-organ alignment panel). We applied a confound battery — registration stress test, blood-core vs wall, lung-air and wall-myocardium partial-volume, tissue density — and a structure/position-matched control (pulmonary artery, deoxygenated, vs aorta, oxygenated). An isotope-matched ^82^Rb uniform-quartz reference bounded the instrument’s positional behaviour. All results were produced by two independent analysis pipelines.

**Results:** RV o-Ps lifetime exceeded LV by Δτ = +0.304 ns (RV 1.700 ± 0.172, LV 1.396 ± 0.130 ns; ~1.4*σ*), in the oxygen-expected direction; the contrast was stable across ± 16 mm registration perturbation (sign preserved in 100% of 342 shifts) and resided in the blood core, not the wall. However, the matched-vessel control was null: pulmonary artery - aorta = -0.011 ± 0.344 ns. Lung-air and wall-myocardium partial-volume were disfavoured, and the effect fell within the isotope-matched ^82^Rb instrumental positional envelope (~0.1–0.35 ns over 40 mm in uniform material).

**Conclusion:** On this single subject, the cardiac o-Ps lifetime contrast does not provide a clean readout of blood oxygenation: an oxygenation effect of the observed (~0.3 ns) magnitude is ruled out by the matched control, while a small physiological effect cannot be excluded. We provide a reusable confound-control battery for evaluating future in-vivo o-Ps oxygenation claims. Multi-subject replication with anatomy decoupled from oxygenation is required.

## 1. Introduction

In a fraction of positron–electron annihilations the pair first forms positronium, a hydrogen-like bound state. Its triplet form, ortho-positronium (o-Ps), is metastable; in matter it localises in nanometre-scale free-volume voids and annihilates predominantly by pick-off, so its lifetime (≈ 1.5–3 ns in soft tissue, versus 142 ns in vacuum) reports the size of those voids and the local electron density. Paramagnetic molecular oxygen further shortens o-Ps lifetime through spin (ortho→para) conversion and oxidation; this oxygen sensitivity is the physical basis for proposing o-Ps lifetime as an imaging biomarker of tissue oxygenation and hypoxia [1,2].

The proposal has gained momentum as positronium lifetime imaging (PLI) has moved from the bench to the clinic. Dedicated multi-photon systems and, more recently, commercial LAFOV PET/CT scanners equipped to register a prompt *γ* in triple coincidence have produced the first in-vivo human positronium images and per-region lifetimes [3–6]. These are real technical achievements. They do not, by themselves, establish that an in-vivo lifetime *contrast* between tissues reflects oxygenation: o-Ps lifetime is dominated by free-volume structure, which varies by ~1 ns between tissue types, whereas the oxygen-specific term implied by published quenching constants is small — on the order of tens of picoseconds across physiological dissolved-oxygen differences [1]. Any in-vivo oxygenation claim therefore faces an identifiability problem, because oxygenation and tissue structure are easily confounded and the oxygen signal is the smaller of the two.

The heart offers an unusually clean natural test. The right ventricle (RV) carries deoxygenated blood and the left ventricle (LV) oxygenated blood; oxygen-quenching predicts a longer o-Ps lifetime in the RV. Crucially, the cardiopulmonary anatomy provides a built-in control that *decouples oxygenation from vessel type and position*: the pulmonary artery carries deoxygenated blood but is, structurally and positionally, a central great vessel like the aorta, which carries oxygenated blood. If a lifetime contrast tracks oxygenation, the pulmonary artery should group with the venous (long-lifetime) side; if it tracks anatomy, the pulmonary artery should group with the aorta. This single comparison breaks the oxygenation/structure degeneracy that an isolated RV>LV difference cannot.

Here we use a public human cardiac PLI dataset to ask whether the RV/LV o-Ps lifetime contrast is attributable to oxygenation. We find a reproducible, registration-stable, blood-borne RV>LV contrast in the oxygen-expected direction — yet the matched-vessel control is null, and the effect lies within the scanner’s own isotope-matched positional envelope. The contribution is twofold: a cautionary single-subject result, and a reusable confound-control battery (matched-vessel test plus exclusion checks plus an isotope-matched instrument reference) for evaluating any future in-vivo o-Ps oxygenation claim.

## 2. Methods

### 2.1 Data

We used publicly deposited, de-identified data from the Bern/Inselspital group acquired on a Biograph Vision Quadra (Siemens Healthineers) [3,4]. The primary subject is an in-vivo [^82^Rb]Cl cardiac acquisition (Zenodo 10.5281/zenodo.11243763): 5.30 × 10^8^ evaluated three-photon events in binary list mode (per event: float64 x, y, z in mm and the prompt→annihilation time difference t in ns), together with the attenuation-correction CT and the reconstructed activity (histo-)image. For instrument characterisation we used the ^82^Rb uniform quartz-glass reference measurement from the same group (Zenodo 10.5281/zenodo.12636019; 3.91 × 10^8^ events). No new data were acquired.

### 2.2 List-mode coordinate mapping and chamber identity

The list-mode axes are not the image axes; we determined the mapping empirically rather than assuming it. Among all 48 axis-permutation/sign combinations, the mapping that reproduces the provider’s own reconstructed activity image — scored by coarse (16-voxel-block, flip-sensitive) correlation — is unambiguous (correlation 0.998; all 47 alternatives *≤* 0.60). Because the CT, activity image and segmentations share an LPS orientation, this fixes RV/LV identity with no flip; the conclusion was cross-checked by a joint multi-organ alignment panel (heart, liver, spleen, both kidneys), which the same mapping wins. No lifetime information enters this determination. (For completeness: the mapping that reproduces the activity image differs from a column-reorder helper distributed with the dataset, which appears to target a CT-aligned frame; the activity-image reproduction is the operative ground truth here.)

### 2.3 Anatomical masks

Organ and chamber masks were generated from the subject’s CT with TotalSegmentator [7] (the total task for organs including liver, spleen and kidneys, and the `heartchambers_highres` task for RV, LV, right and left atria, aorta, pulmonary artery and myocardium).

### 2.4 Lifetime model

For each region we accumulated a Δ*t* histogram (0.1 ns bins) and fitted a background plus a two-component model: a flat accidental background B, a short component, and the o-Ps component, each a decay exponential convolved with a Gaussian instrument response (an EMG), over t *∈* [-3, 12] ns with Poisson weights. τ_oPs_ is the long component. A model-independent tail single-exponential fit (after the prompt peak) served as a cross-check; it agreed on sign and approximate magnitude. The same model was applied identically to every region, so model systematics partially cancel in the *contrast*.

### 2.5 Registration stress test

We modelled registration uncertainty as a global integer-voxel translation of the event cloud relative to the masks (equivalently, shifting both masks together), and refitted every shift. Two sweeps were run: a dense ± 3-voxel grid (*≤* 8 mm, 342 shifts) and a per-axis sweep to ± 10 voxels (± 16 mm).

### 2.6 Confound battery

We tested, individually: (i) chamber-wall partial volume, by eroding masks to their blood core versus the near-wall rim; (ii) lung-air partial volume, by correlating τ with a lung-air fraction field (lung-lobe masks convolved with the PET point-spread function); (iii) wall-myocardium leakage, by correlating τ with a myocardium point-spread field; (iv) tissue density, via per-compartment CT Hounsfield units; and (v) instrument/reconstruction positional behaviour, using the isotope-matched ^82^Rb uniform-quartz reference, in which any spatial lifetime variation is by construction non-biological.

### 2.7 Reproducibility

The analysis was implemented and executed by two independent pipelines that converged on every reported number; during development the second pipeline twice identified and corrected an over-statement (see Declarations and the public code’s audit log). Software: Python 3.10, NumPy, SciPy, NiBabel; scripts and intermediate products are released (Section: Code availability).

## 3 Results

### 3.1 A reproducible, registration-stable RV>LV contrast

Per-chamber lifetimes were τ_oPs_(RV) = 1.700 ± 0.172 ns (n = 2.36 × 10^6^ events) and τ_oPs_(LV) = 1.396 ± 0.130 ns (n = 1.95 × 10^6^), giving **RV - LV = +0.304 ns (about 1.4sigma20 20 12 61 79 80 81 98 701 33 100 204 250 395 398 399 400** — the direction oxygen-quenching predicts (Figure 1). The contrast was robust to registration: across the dense ± 3-voxel grid the sign was preserved in 100% of 342 shifts with a dispersion of only 0.09 ns, and it remained positive along every single axis out to ± 16 mm (Figures 2–4). Registration is therefore not the limiting factor.

**Figure 1.**
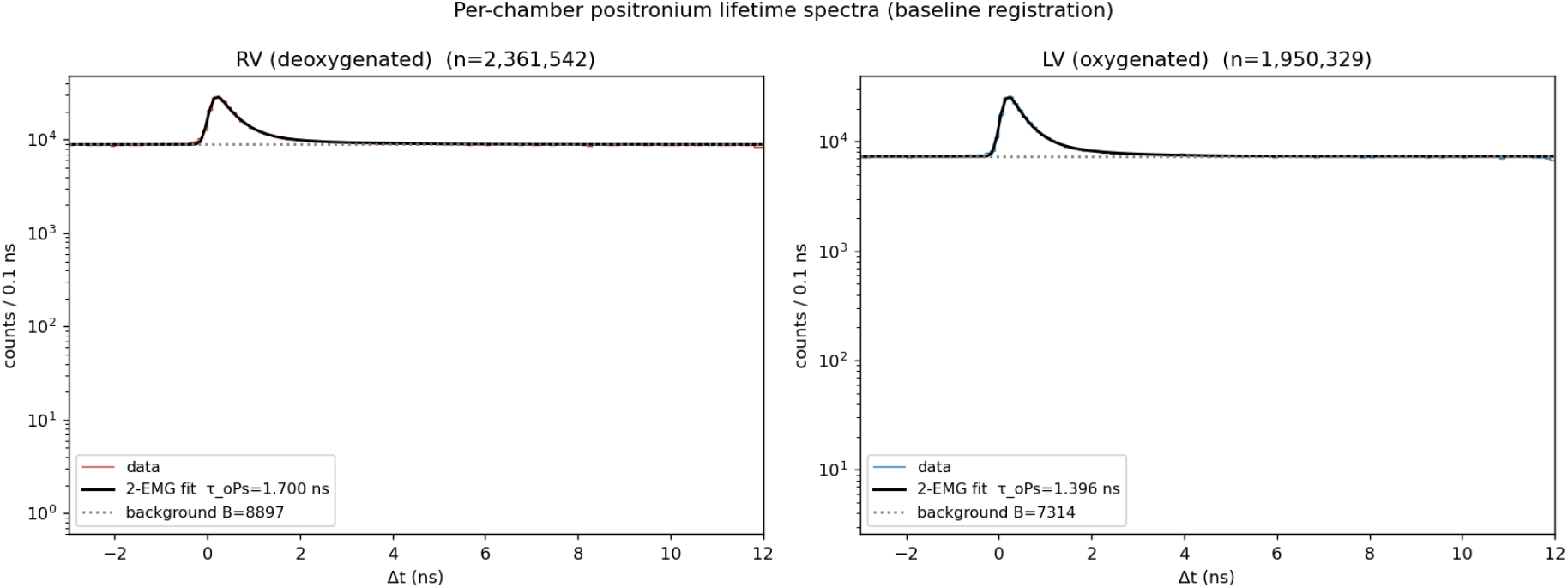
Per-chamber positronium-lifetime spectra (Δ*t* histograms, log scale) for the right and left ventricles at baseline registration, with the background-plus-two-component EMG fits and the fitted flat background. The o-Ps signal is a modest excess over a large accidental background; τ_oPs_(RV) = 1.700 ns, τ_oPs_(LV) = 1.396 ns.

**Figure 2.**
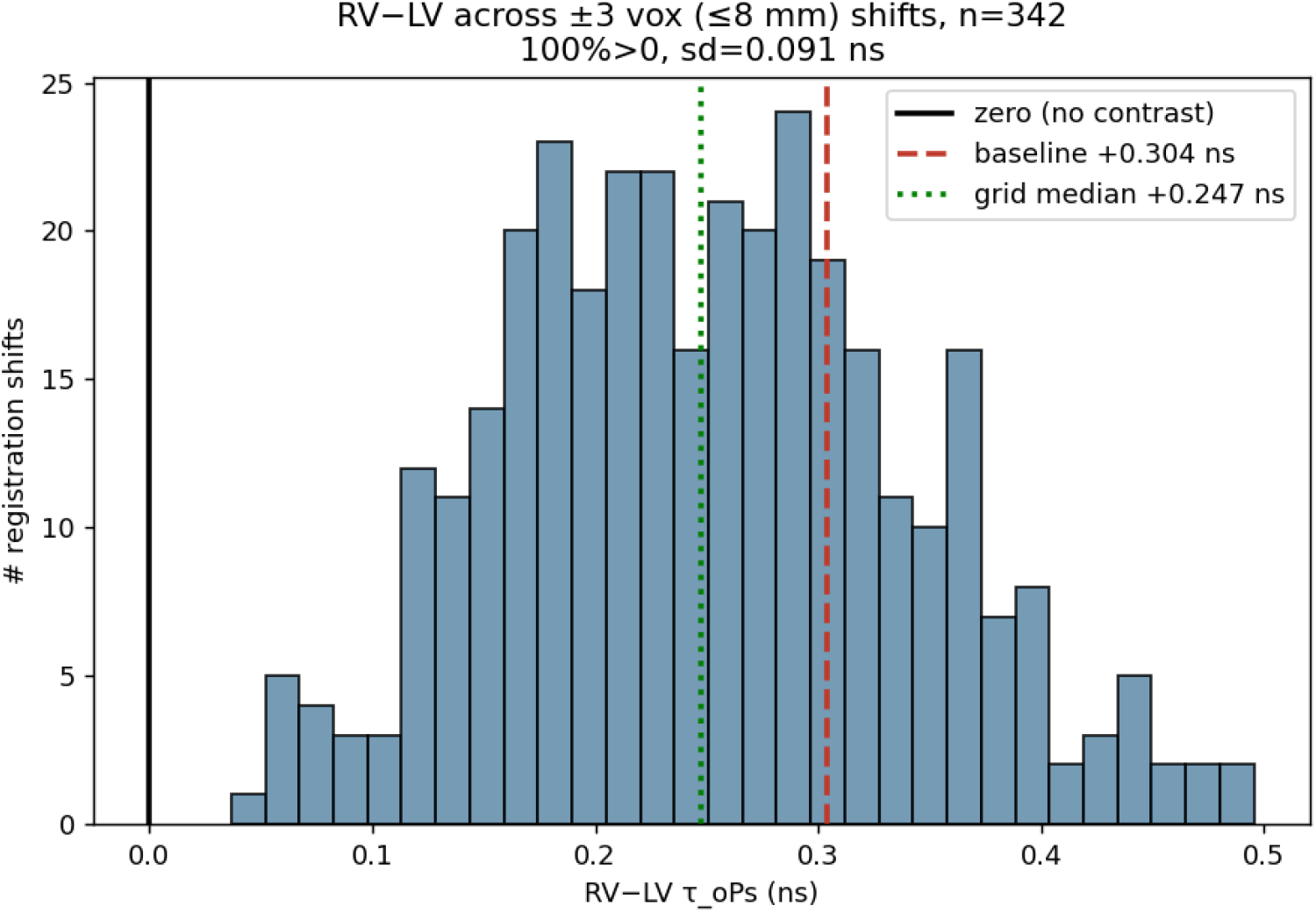
Distribution of the RV-LV lifetime difference across the dense ± 3-voxel (*≤* 8 mm) registration grid (n = 342 shifts). The contrast is positive in 100% of shifts (dispersion 0.09 ns); dashed line, baseline; the difference never crosses zero.

**Figure 3.**
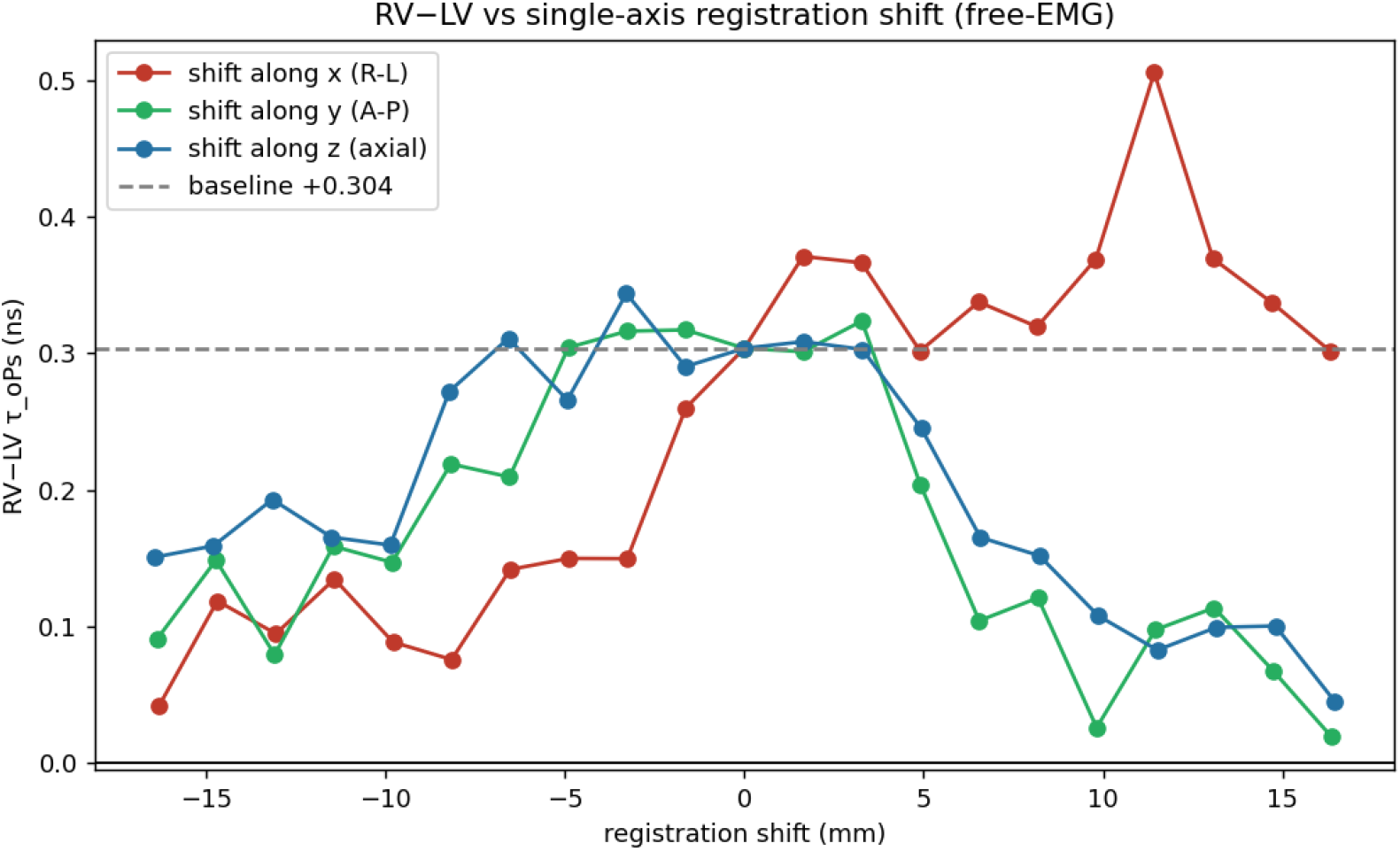
RV-LV lifetime difference versus single-axis registration shift (± 16 mm) along each axis. The difference stays positive across the full tested range.

**Figure 4.**
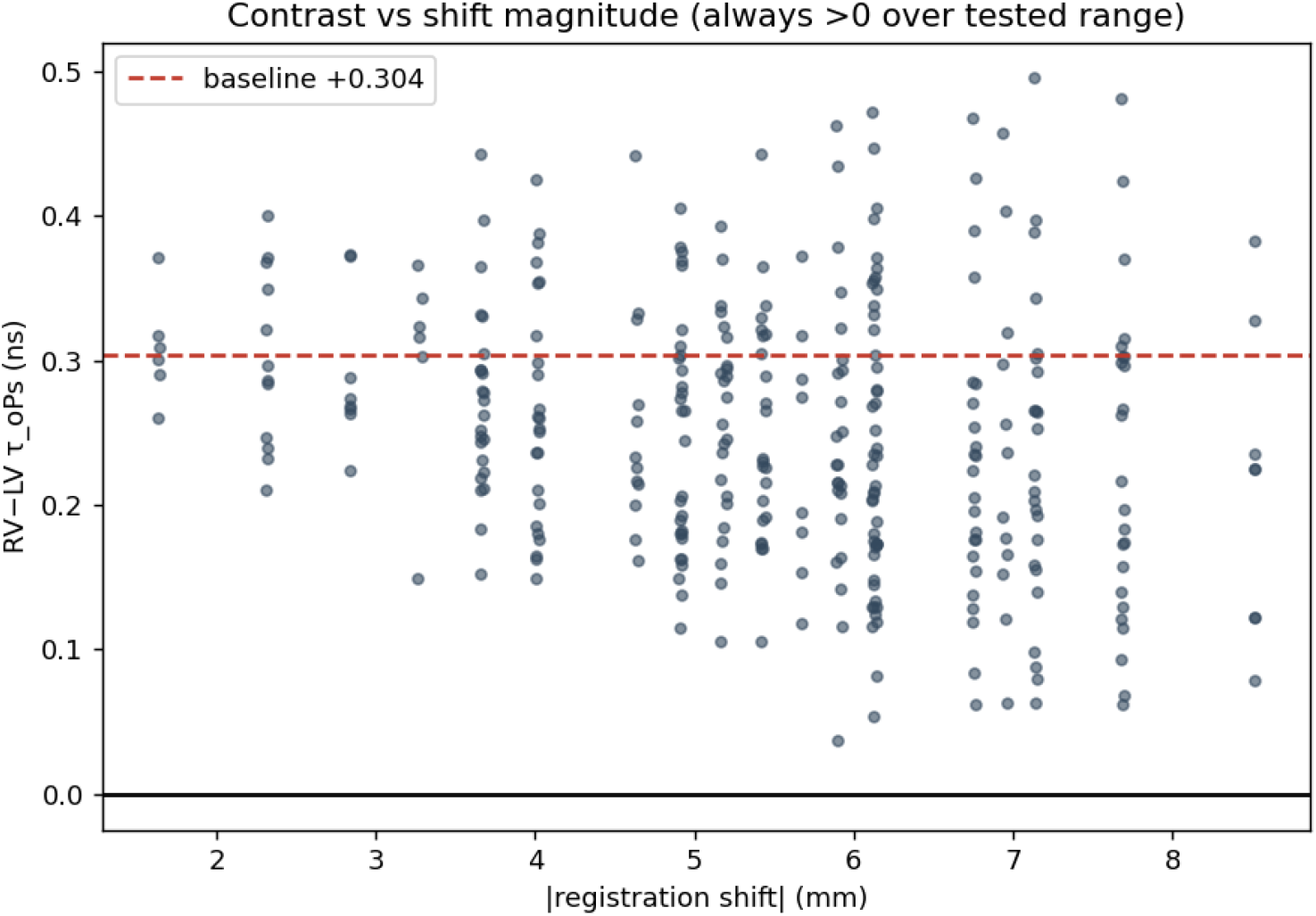
RV-LV lifetime difference versus the magnitude of the registration shift; positive throughout.

### 3.2 The contrast resides in blood, not the wall

Eroding each chamber to its blood core *increased* the contrast (core Δτ = +0.43 ns), whereas the near-wall rim erased it (RV rim 1.54 ≈ LV rim 1.51 ns, Δτ = +0.03 ns). Partial volume from the chamber wall was diluting, not creating, the contrast.

### 3.3 Chamber identity is confirmed

The mapping underlying these assignments reproduces the provider’s activity image at correlation 0.998 (versus *≤* 0.60 for all alternative orientations) and wins the joint multi-organ alignment panel (Section 2.2). RV is anatomically the right ventricle; there is no flip. This was established without using any lifetime information.

### 3.4 Oxygenation ladder and the matched-vessel control (key result)

Extending to the cardiopulmonary compartments (Table 1, Figure 5) shows that the venous>arterial tendency is carried by the right-heart chambers, not by oxygenation per se. The decisive comparison is the structure- and position-matched control: the **pulmonary artery (deoxygenated) and the aorta (oxygenated), two central great vessels, have indistinguishable lifetimes — pulmonary artery - aorta = -0.011** ± **0.344 ns**. The ± 0.34 ns uncertainty exceeds the dissolved-oxygen effect expected on physiological grounds (tens of ps), so this control rules out an oxygenation effect of the RV/LV *magnitude* (~0.3 ns) but does not exclude a small physiological effect.

**Table 1.** Per-compartment o-Ps lifetime (eroded blood cores).

| Compartment | Class | $\tau_{\text{oPs}}$ (ns) | n ($\times 10^6$) |
| --- | --- | --- | --- |
| Right ventricle | deoxygenated | $1.731 \pm 0.197$ | 2.05 |
| Right atrium | deoxygenated | $1.640 \pm 0.332$ | 1.25 |
| Superior vena cava | deoxygenated | $1.444 \pm 0.505$ | 0.30 |
| Inferior vena cava | deoxygenated | $1.425 \pm 0.412$ | 0.33 |
| **Pulmonary artery** | **deoxygenated** | **$1.259 \pm 0.312$** | 0.54 |
| Left atrium | oxygenated | $1.302 \pm 0.270$ | 0.90 |
| Left ventricle | oxygenated | $1.358 \pm 0.134$ | 1.71 |
| **Aorta** | **oxygenated** | **$1.270 \pm 0.145$** | 1.74 |
| Myocardium (control) | muscle | $1.662 \pm 0.206$ | 1.10 |

**Table 2.** Confound-control battery.

| Confound | Test | Result |
| --- | --- | --- |
| Registration error | translational sweep $\pm 16$ mm | sign-stable (100%/342 shifts, $\sigma$ 0.09 ns) — not the driver |
| Chamber-wall partial volume | core vs near-wall rim | core $\Delta +0.43$; rim $\Delta +0.03$ — signal is in blood, not wall |
| **Blood oxygenation** | matched control (pulmonary artery vs aorta) | $\Delta = -0.01 \pm 0.34$ ns — no effect of the RV/LV magnitude |
| Lung-air partial volume | lung-PSF field vs $\tau$ | air $\approx 0$ in cores; r = -0.04 — disfavoured |
| Wall-myocardium leakage | myocardium-PSF field vs $\tau$ | LV has $3 \times$ more yet lower $\tau$; r = +0.06 — refuted |
| Instrument/reconstruction position | isotope-matched $^{82}\text{Rb}$ uniform quartz | positional envelope $\sim 0.1\text{--}0.35$ ns / 40 mm — effect lies within it |
| Tissue density | per-compartment CT HU | blood/muscle 86% overlap — uninformative |

**Figure 5.**
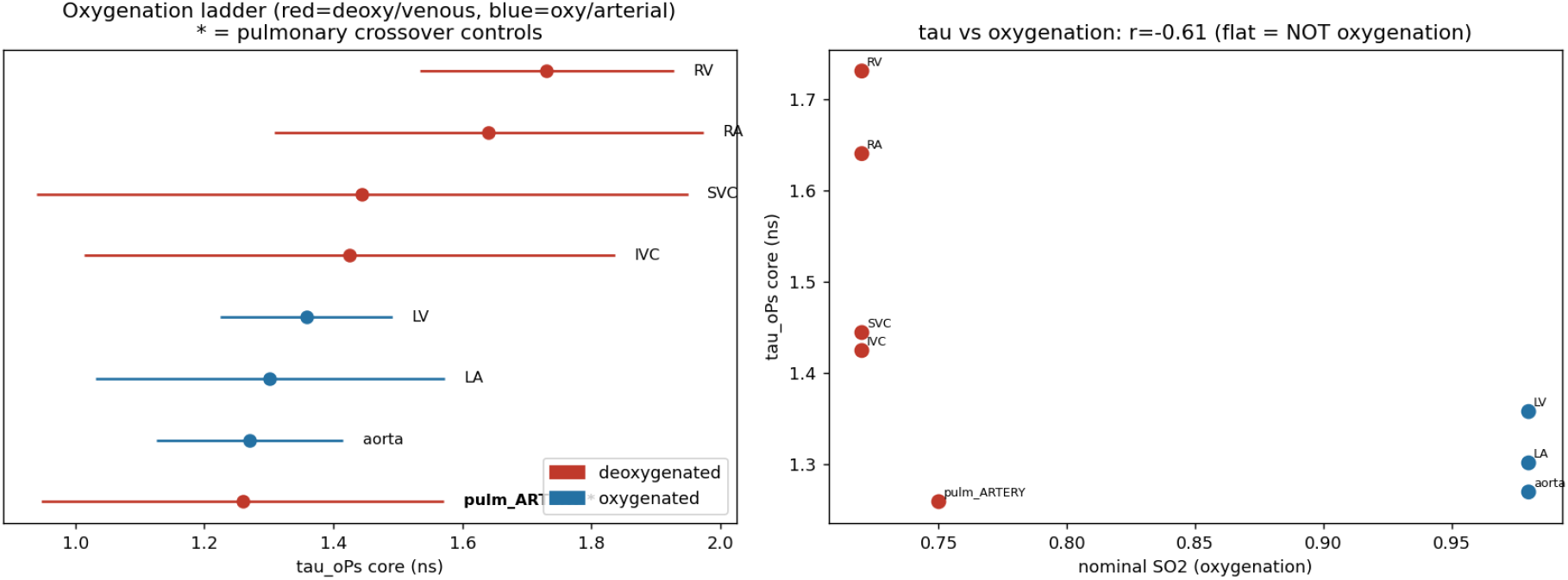
Oxygenation ladder. (Left) per-compartment o-Ps lifetime, coloured by oxygenation class, with the pulmonary crossover controls highlighted; (right) lifetime versus nominal oxygen saturation. The flat trend and the null pulmonary-artery–aorta pair indicate the contrast does not track oxygenation.

**Figure 6.**
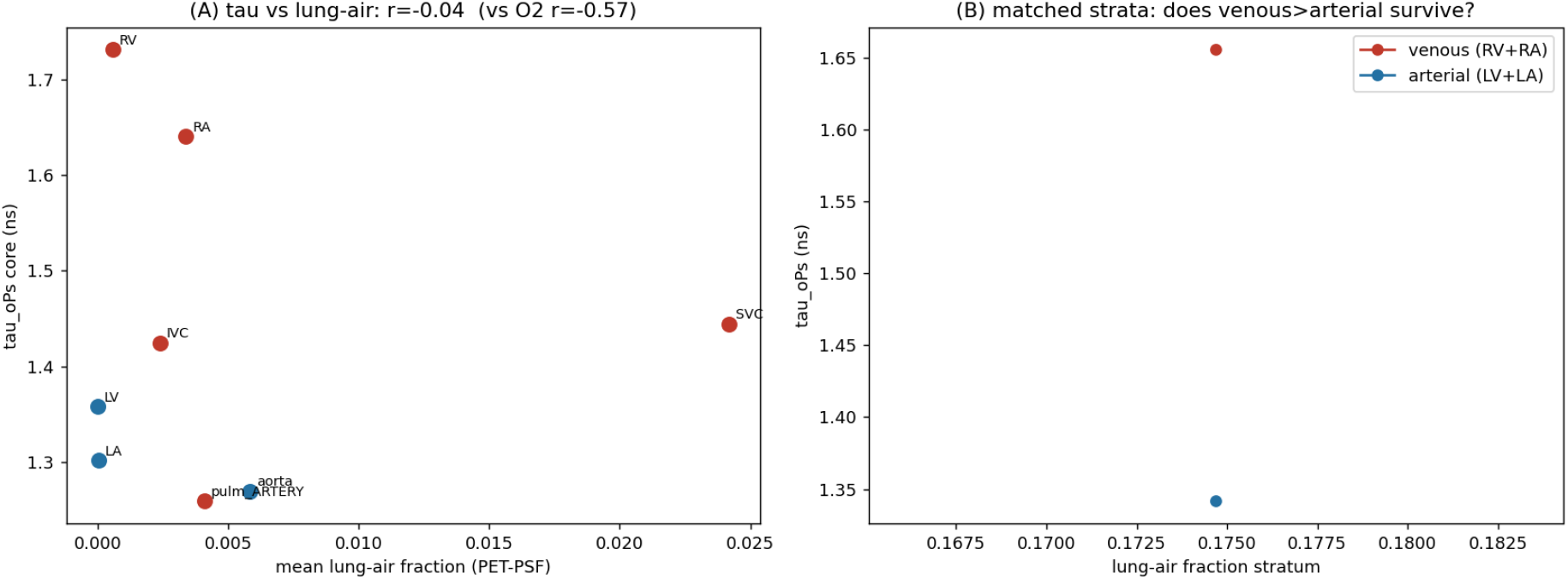
Lung-air partial-volume check: o-Ps lifetime versus the local lung-air fraction (and the matched-stratum comparison). The blood cores are essentially air-free and lifetime does not track the lung-air fraction.

### 3.5 Confound exclusions

Lung-air partial volume is disfavoured (the eroded blood cores are essentially air-free and τ does not correlate with the lung-air fraction, r = -0.04). Wall-myocardium leakage is refuted directly: the LV core has ~3 × more nearby myocardium than the RV core (myocardium point-spread 0.044 vs 0.014) yet a *lower* lifetime (r(τ, myocardium-PSF) = +0.06). CT Hounsfield units do not separate blood from muscle (86% overlap) and are uninformative.

### 3.6 Isotope-matched instrumental positional envelope

In the uniform ^82^Rb quartz reference — where any spatial lifetime variation is non-biological — the instrument response is consistent with the human fits (prompt offset t0 = 0.067 ns ≈ human 0.066; Gaussian width *σ* = 0.088 ns) and the absolute lifetime is reasonable (1.71 ns versus 1.589 ns by the original Bayesian/voxel-wise method; the difference reflects model and volume-of-interest choices). Critically, the reconstructed lifetime in this single uniform material **varies by ~0.1–0.35 ns over ~40 mm** (a systematic envelope, not a calibrated gradient — its direction is unstable across core sizes). This isotope-matched envelope is comparable to the entire RV-LV effect; the previously cited ^124^I phantom bound (*≤* 0.08 ns) understates it, plausibly because ^82^Rb’s high positron energy degrades spatial localisation. The cardiac contrast therefore falls within the scanner’s own positional behaviour.

### 3.7 Residual right-heart elevation: mechanism open

After excluding oxygenation (of the observed magnitude), lung-air, wall-myocardium, registration and gross instrumental bias, what remains is a ~1.5*σ* elevation specific to the right-heart chambers whose mechanism we leave open. An interim hypothesis (internal trabecular muscle plus ^82^Rb myocardial avidity) is plausible but unproven; regional scatter and bolus kinetics are not excluded.

## 4. Discussion

The natural reading of an isolated RV>LV lifetime difference in the oxygen-expected direction is that o-Ps lifetime is sensing blood oxygenation. Three lines of evidence argue against that reading here. First, the magnitude (~0.3 ns) is roughly an order of magnitude larger than the dissolved-oxygen effect implied by published quenching constants for the arterial–venous difference [1] — so, if real, it is too large to be dissolved-oxygen quenching alone. Second, and decisively, the structure- and position-matched control (pulmonary artery versus aorta) is null: when oxygenation is varied while structure and position are held fixed, no lifetime difference appears. Third, the apparent venous>arterial trend is carried by the right-heart chambers, which differ from the great vessels in trabeculation, wall geometry and surrounding tissue, and the effect lies within the scanner’s own isotope-matched positional envelope.

Why is the contrast so easily confounded? In cardiac anatomy, “deoxygenated” travels with “right heart / specific position / specific structure”, so a single RV>LV comparison cannot separate oxygenation from anatomy. The pulmonary vessels are the one place where oxygenation and vessel type disagree, which is exactly why the pulmonary-artery-versus-aorta control is informative. We also note that deoxyhaemoglobin, the dominant paramagnetic species in venous blood, would shorten the RV lifetime — the opposite of what is observed — so it is not driving the contrast either. Finally, ^82^Rb is a high-positron-energy, myocardium-avid tracer; both properties (degraded localisation and muscle uptake) work against a clean blood-pool oxygenation read.

The practical implication for in-vivo PLI is methodological: a lifetime contrast between two anatomical regions is not evidence of an oxygenation difference unless oxygenation has been varied with structure and position controlled, and unless the contrast exceeds the scanner’s isotope-matched positional envelope at adequate effective statistics. The confound-control battery used here — a matched-vessel control, blood-core isolation, partial-volume checks, and an isotope-matched uniform-material reference — is portable to any future o-Ps oxygenation claim and is released with this work.

## 5. Limitations

This is a single subject, and the central contrasts are weak (~1.5*σ*): the robust findings are the *no-detectable-difference* of the matched control and the disfavouring of named confounds, not a positive claim of zero oxygenation effect. The matched-control uncertainty (± 0.34 ns) exceeds the expected physiological dissolved-oxygen effect (tens of ps), so a small oxygenation contribution cannot be excluded. The instrumental positional envelope is a systematic bound, not a calibrated gradient (its direction is unstable across analysis volumes). The o-Ps component sits on a large flat accidental background, so its effective statistics are modest, and the absolute lifetime is model-dependent (~0.1 ns; our quartz value 1.71 vs the reference 1.589 ns). Masks were derived from the subject’s CT; thin systemic vessels were too noisy to use. The mechanism of the residual right-heart elevation is unresolved. The strongest version of this test requires replication across subjects with anatomy decoupled from oxygenation, and, ultimately, an independent oxygenation ground truth.

## 6. Conclusion

In a public human cardiac LAFOV PET/CT dataset, a reproducible, registration-stable, blood-borne and chamber-identity-confirmed RV>LV o-Ps lifetime contrast (~0.3 ns) in the oxygen-expected direction is **not attributable to blood oxygenation at the achievable precision**: the structure/position-matched pulmonary-artery-versus-aorta control is null, and the effect lies within the scanner’s own isotope-matched positional envelope. In-vivo o-Ps lifetime did not yield a clean oxygenation readout here. We release a reusable confound-control methodology to evaluate such claims, and call for multi-subject, oxygenation-decoupled replication before cardiac o-Ps lifetime contrasts are interpreted as oxygenation maps.

## Data Availability

All input data are publicly available on Zenodo. The human [82Rb]Cl PET/CT dataset is available at https://doi.org/10.5281/zenodo.11243763
and the 82Rb quartz reference dataset is available at https://doi.org/10.5281/zenodo.12636019.
No raw data are redistributed in this manuscript. Analysis code, result tables, figures, and reproduction instructions are available at https://github.com/Jefemaestro33/rvlv-paper.

https://doi.org/10.5281/zenodo.12636019

https://github.com/Jefemaestro33/rvlv-paper

## Data Availability

https://doi.org/10.5281/zenodo.12636019

https://github.com/Jefemaestro33/rvlv-paper

## Declarations

### Ethics approval and consent

This work is a secondary analysis of publicly available, fully de-identified data; it involved no new human-subjects data collection and required no additional ethical approval. The original invivo data were acquired and shared by the Bern/Inselspital group under their institutional approvals [3,4] and are distributed under CC-BY-4.0; the analysed CT does not include the face. The author had no access to identifiable participant information.

### Data availability

All inputs are public on Zenodo: the [^82^Rb]Cl human dataset (10.5281/zenodo.11243763) and the ^82^Rb quartz reference (10.5281/zenodo.12636019); a ^124^I phantom record (10.5281/zenodo.13443797) is referenced for context. No raw data are redistributed here.

### Code availability

Analysis code, intermediate result tables, figures, a key-numbers file, environment specification and step-by-step reproduction instructions are available at https://github.com/Jefemaestro33/rvlv-paper (MIT for code; CC-BY-4.0 for text and figures). An archival DOI can be added after public release.

### Competing interests

The author declares no competing interests.

### Funding

No specific funding was received for this work.

### Author contributions

E.D.P.Z. conceived the study, performed the analysis, and wrote the manuscript. Analyses were implemented and independently cross-checked by two AI-assisted pipelines under the author’s direction; the AI systems are tools and are not authors.

## Acknowledgements

We thank L. Mercolli, W. Steinberger and colleagues (Inselspital Bern) for openly depositing the positronium-lifetime data that made this re-analysis possible.

## Notes

### Competing Interest Statement

The authors have declared no competing interest.

### Author Declarations

The study used only openly available human data that were available before the initiation of this secondary analysis. The source human PET/CT data were deposited publicly by the Bern/Inselspital group on Zenodo and can be located at https://doi.org/10.5281/zenodo.11243763. The isotope-matched 82Rb quartz reference dataset used for instrument characterization was also publicly available on Zenodo before the initiation of this study and can be located at https://doi.org/10.5281/zenodo.12636019. No restricted-access, application-based, or non-public human data were used.

